# Trends in diagnostic tests ordered for children: a retrospective analysis of 2 million laboratory test requests in Oxfordshire, UK from 2005 to 2019

**DOI:** 10.1101/2022.12.15.22283423

**Authors:** Elizabeth T Thomas, Diana R Withrow, Brian Shine, Peter J Gill, Rafael Perera, Carl Heneghan

**Affiliations:** Centre for Evidence-Based Medicine, Nuffield Department of Primary Care Health Sciences, Radcliffe Observatory Quarter, University of Oxford, Oxford, United Kingdom; Nuffield Department of Primary Care Health Sciences, Radcliffe Observatory Quarter, University of Oxford, United Kingdom; Department of Clinical Biochemistry, John Radcliffe Hospital, Oxford, United Kingdom; Department of Paediatrics, Faculty of Medicine, University of Toronto, Toronto, Ontario, Canada; Hospital for Sick Children, Toronto, Ontario, Canada

## Abstract

**Summary:** Diagnostic tests play an essential role in children’s health. Previous work has shown variation in the use of diagnostic tests for adults. However, comparatively little is known about the use of tests in children. We aimed to analyze temporal trends in laboratory testing for children aged 0 to 15 from 2005 to 2019 in Oxfordshire, United Kingdom.

**Methods:** For this retrospective analysis, we used data from the Oxfordshire University Hospital NHS Trust laboratories. Using joinpoint regression models, we estimated annual percentage changes (APC) in test use. Temporal changes in age-adjusted rates in test use were calculated overall and stratified by healthcare setting, sex and age.

**Findings:** Between 2005 and 2019, overall test use increased in children (APC 1.6%, 95 confidence interval -0.8% to 4.1%). Increases were highest in females, in those aged 11-15 years and the outpatient setting. The most frequently requested tests were full blood count, urea and electrolytes, liver function test, C-reactive protein and calcium magnesium phosphate. The test with the greatest increase in use was Vitamin D, which increased on average by 27% per year. Other tests that showed a significant temporal increase included parathyroid hormone, iron studies, folate, vitamin B12 levels, glucose, HbA1c, IgA, coeliac, creatine kinase, thyroid function tests and IgG/IgM. Test changes were not uniformly distributed across all settings and age groups.

**Interpretation:** The increase in test use may be the result of a combination of factors, including changes to the health service resulting in an increased volume of presentations and referrals, shifts in workforce composition towards less experienced clinicians, increased parental anxiety and expectation of tests and/or increased awareness and changing prevalence of disease. Further research is needed to quantify whether test use is warranted and to compare trends in Oxfordshire with other settings.

**Funding:** No funding was obtained for this study.

**Research in context:** *Evidence before this study:* We searched PubMed using the terms “diagnostic test” “child” and “variation” from inception until 7 November 2022 to identify studies related to diagnostic test use in children. Previous studies have demonstrated substantial variation in the use of diagnostic tests across primary and secondary care in the UK. However, most of the literature on diagnostic testing focused on adults. Population-based studies of UK primary care identified that test use had increased by 9% annually from 2000 to 2015. Tests with the highest increase were knee MRIs which increased by 69% per year, followed by vitamin D tests and brain MRI. Tests subject to the greatest practice variation included drug monitoring, urine microalbumin and pelvic CTs. However, these studies did not specifically analyse data on test use in children. A few studies on children examined variation in the use of tests for specific conditions, such as community-acquired pneumonia, orbital cellulitis, fever or diabetes. No studies have quantified test use across all settings or examined temporal trends in test use in children.

*Added value of this study:* To our knowledge, this is the first population-based study to estimate long-term trends in childhood test use. We collected data on laboratory tests that were analysed at the Oxfordshire University Hospital NHS Trust laboratories for children aged below 16 between 2005 and 2019 and evaluated average annual percentage change and annual percentage change in test use using joinpoint regression. We found that test use increased by 2% per year overall, with the highest increases in the outpatient setting and for females aged 11-15. Vitamin D tests experienced the greatest overall increase during the study period.

*Implications of all the available evidence:* Changes in test use may suggest potentially inappropriate testing, especially for Vitamin D and C reactive protein. It also suggests an increase in disease awareness and prevalence as well as changes in the healthcare workforce and service provision. A comparison between testing rates and the corresponding test results must be made to better understand whether increased test use is warranted. The observed trends in this study should also be compared with other settings to determine their generalisability. We encourage clinicians to become aware of their test-ordering practices and consider the individual and systemic implications of testing in children.

## Background

Diagnostic testing plays an important role in the provision of health care. In England, laboratory and pathology services (including biochemistry, haematology, microbiology, histopathology and cytology tests) are estimated to cost 2·5 billion pounds annually, comprising 3-4% of the NHS budget.^1^ Similar data from the United States suggests that expenditure on diagnostic testing was 82·7 billion dollars in 2017, or 2% of total US healthcare spending.^2^ It is estimated that 70 – 80% of all healthcare decisions affecting diagnosis or treatment involve a pathology test.^1,3,4^ Between 2004 and 2006 in the UK, demand for pathology services increased at an annual rate of 10%.^1^

Substantial variation has been demonstrated in diagnostic test use across primary and secondary care in the UK.^5–7^ The 2017 Atlas of Variation in NHS Diagnostic explored unwarranted variation in a range of imaging, endoscopy, physiological and screening services. However, most of the reported diagnostic measures focused on adults, as does much of the existing literature on diagnostic testing.^7,8^ Children constitute 19% of the UK’s population^9^ and over 85% of children are registered with an NHS general practitioner (GP), who act as gatekeepers to paediatric specialist care.^10,11^ Clinicians often face uncertainty when deciding whether to order diagnostic investigations for the children as failure to perform necessary diagnostic laboratory tests can lead to missed diagnoses but unnecessary diagnostic laboratory tests (over-testing) may lead to physical and psychological harms to children as well as straining already limited health care resources.^12^ Parents are also reluctant to obtain blood samples from children given the distress it causes.

There is a paucity of comprehensive data that explores laboratory testing in children. This study aims to determine the most frequently performed diagnostic laboratory tests for children in the Oxfordshire region from 2005 to 2019, the temporal variation in these tests and the proportion of tests ordered in primary care compared with secondary care.

## Methods

### Study design and Data sources

This was a retrospective observational study of diagnostic laboratory test data.

### Setting

We obtained laboratory testing data from Oxford University Hospitals and Oxfordshire General Practices from 1^st^ January 2005 to 31^st^ December 2019. This was selected as the study end date to eliminate the impacts of the COVID-19 pandemic. The laboratory is the sole referral centre for 67 general practices and four hospitals, making up over 95% of the tests carried out in the county. Data from all laboratory tests conducted among children aged 0 to 15 were included. We excluded point-of-care tests such as blood gas and glucose tests as, in practice, these can be performed at the bedside and are not consistently sent to the laboratory for analysis.

### Variables and data sources

The Oxford University Hospitals (OUH) Trust database contains previously collected laboratory test data. We extracted non-identifiable data, including the name of the test, indication for the test, patient sex and age, and whether the test was ordered in primary or secondary care. The test codes for each test panel are provided in the **Supplementary File**.

### Statistical analysis

We estimated the proportion of tests requested in general practice, inpatient and outpatient (hospital paediatric clinic) settings each year. We estimated crude and age-standardised test rates per 1,000 child years using the 2019 population as the standard. Testing rates were stratified by gender and age; under 1 year (infants); 1-5 years (early childhood); 6-10 years (middle childhood), and 11-15 years (adolescence) (12).

We used joinpoint regression to model temporal changes in age-adjusted rates from 2005 to 2019. Points where significant changes in rates occurred (joinpoints) were identified and annual percentage changes (APC) between joinpoints were estimated. We also estimated the average annual percentage change (AAPC), a summary measure of the trend from 2005 to 2019, stratified by setting, sex, and age. APCs and AAPCs were estimated for the 25 most frequently requested tests. APCs and AAPCs were modelled in Joinpoint software, and all other statistical analyses were performed using R.

### Ethics Approval

The University of Oxford Clinical Trials and Research Governance committee waived ethical approval for this work. A data protection impact assessment was conducted and approved.

## Results

### Characteristics of included participants

There were 1,749,425 tests performed on 113,607 children from 1 January 2005 to 31 December 2019, of which 46% (52,207 of 113,607) were females. 71% of tests (1,232,556 of 1,749,425) occurred in the inpatient setting, 17% in general practice, and 13% in the outpatient setting. Children had a median of five tests each (IQR 3 to 8). The median number of tests per child for each setting is shown in Table 1. One-third of tests (33%, n= 580,636) were performed in the under-1 age group, of which most (96%, n=558,716) were performed in the inpatient setting (**Supplementary Table 1**).

**Table 1.** Characteristics of included patients and tests.

|  | Number of children | % |
| --- | --- | --- |
| Females | 52,207 | 46.0 |
| Males | 61,400 | 54.0 |
| Total | 113,607 | 100 |
| Age group | Number of tests | % |
| Total tests | 1,749,425 | 100.0 |
| <1 year | 580,636 | 33.2 |
| 1-5 years | 439,770 | 25.1 |
| 6-10 years | 319,387 | 18.3 |
| 11-15 years | 409,632 | 23.4 |
| Setting |  |  |
| GP | 293,506 | 16.8 |
| IP | 1,232,556 | 70.5 |
| OP | 223,363 | 12.8 |
|  | Median no. of tests per child* | Interquartile Range |
| Total | 5 | 3, 8 |
| GP | 5 | 3, 8 |
| IP | 5 | 3, 7 |
| OP | 4 | 2, 8 |

\*In children who had at least one test

### Temporal change in test use

The age-adjusted rate of total test use increased from 878 tests per 1,000 child years in 2005 to 1,107 tests per 1,000 child years in 2019 (**Figure 1a** AAPC 1·5% [95% CI -0·8 to 3·9%, p=0·2]). This net increase occurred despite an initial decrease of 2·2% per year between 2005 and 2012 (95% CI -0·6 to -3·8%, p=0·01, Figure 1). From 2012 to 2015, the APC increased to 9·0% per year (95% CI -3·0 to 22·5%) and then between 2015 and 2019 was 2·8% per year (95% CI -0·5 to 6·3%).

**Figure 1.**
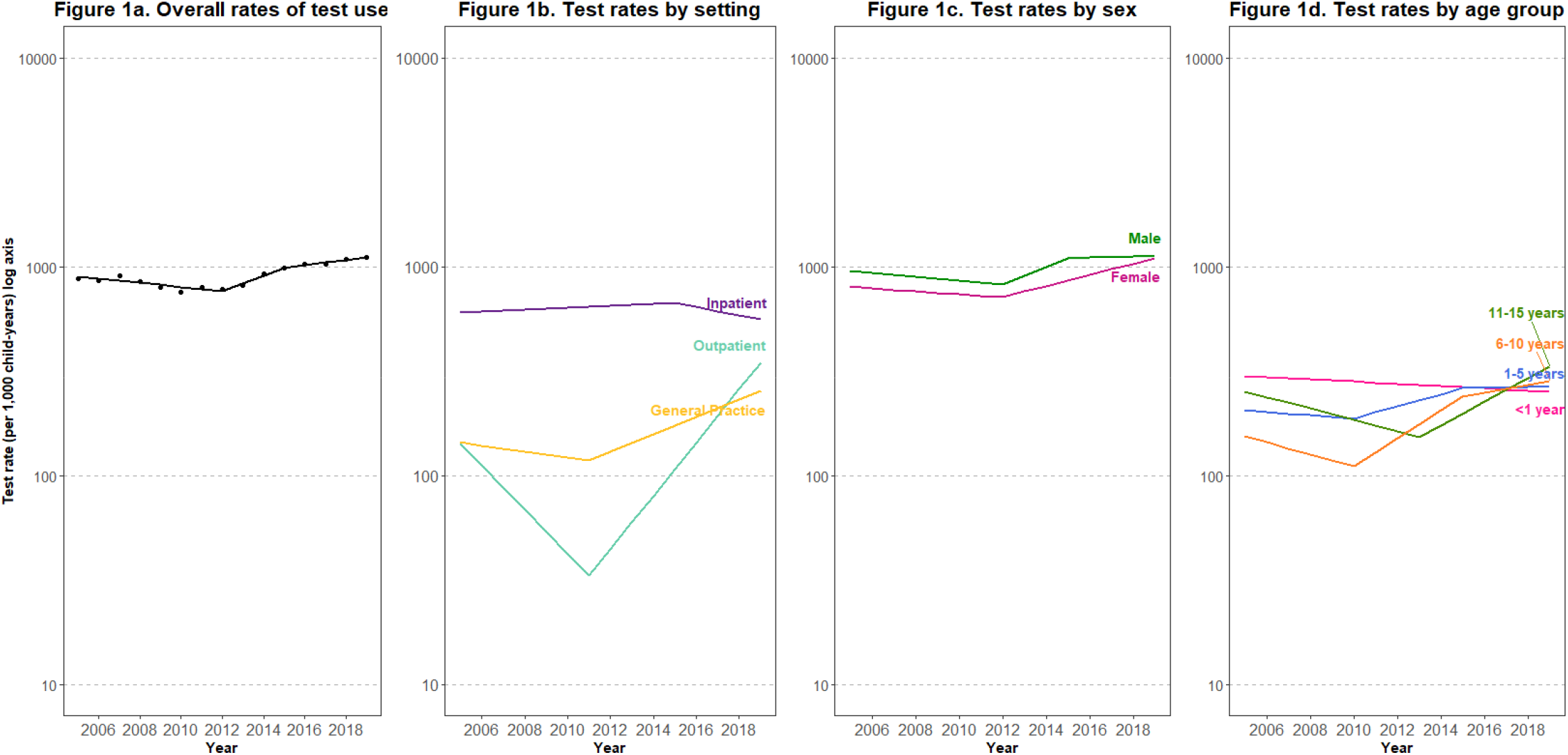
Test use among children in Oxfordshire from 2005 to 2019. 1a) Overall test use 1b) By health care setting 1c) By sex 1d) By age group. **Footnotes:** AAPC: Average annual percentage change; APC: annual percentage change Figure 1a: AAPC = 1.5% (95% CI -0.8 to 3.9%, p=0.2) APC 2005 – 2012 = -2.2% (95% CI -3.8 to -0.6%, p=0.01) APC 2012 – 2015 = 9.0% (95% CI -3.0 to 22.5%, p=0.1) APC 2015 – 2019 = 2.8% (95% CI -0.5 to 6.3%, p=0.1) 1b: AAPC by setting. General Practice = 4.2% (95% CI 1.5 to 6.9%, p= 0.002) Inpatient = -0.6% (95% CI -2.1 to 0.9%, p=0.4) Outpatient = 6.6% (95% CI 1.9 to 11.5%, p=0.005) 1c: AAPC by sex. Female = 2.3% (95% CI 1.3 to 3.3%, p <0.001) Male = 1.2% (95% CI -1.3 to 3.8%, p=0.3) 1d: AAPC by age group. <1 year = -1.2% (95% CI -2.2 to -0.2%, p=0.02); 1-5 years= 1.9% (95% CI -0.8 to 4.6%, p=0.2); 6-10 years = 4.4% (95% CI 2.6 to 6.3%, p <0.001) 11-15 years = 2.0% (95% CI -1.1 to 5.2%, p =0.2)

Figure 1b shows a temporal change in test use by setting. Test use decreased overall in the inpatient setting. In general practice, test use initially declined by 3·3% per year until 2011 (95% CI -8·3 to 1·9%, p=0·2) and then increased by 10·1% per year (95% CI 6·5 to 14·0%, p<0·001). Testing in outpatients followed a similar trend, with testing initially decreasing by 21.6% per year until 2011 (95% CI -28·3 to -14·2%, p<0.001) and then sharply increasing by 34% per year (95% CI 26·7 to 42·2%, p<0·001).

Figure 1c illustrates test use by sex. Testing rates for males and females followed similar trends until 2015 when test use in males stabilized, growing by 1·0% per year (95% CI -2·6 to 4·7%, p=0·5), whereas testing in females continued to rise by 6·4% per year from 2012 (95% CI -4·7 to 8·0%, p<0·001).

The rates of test use by age group are presented in Figure 1d. Test use declined overall in children under 1 year. Testing in all other age groups increased; this was particularly striking for children aged 11-15 years, where testing increased by 13.8% per year from 2013 (95% CI 6·9 to 21·1%, p<0·001).

The proportion of children in Oxfordshire receiving at least one test in any setting increased by 39% (from 8·8% in 2005 to 12·3% in 2019, see **Figure 2**). In the outpatient setting, the proportion of children receiving at least one test increased by 84% (from 2·2% to 4·0%). In the inpatient setting this increased by 30% (from 4·7 to 6·1%) and in general practice by 42% (2·2 to 4·0%).

**Figure 2.**
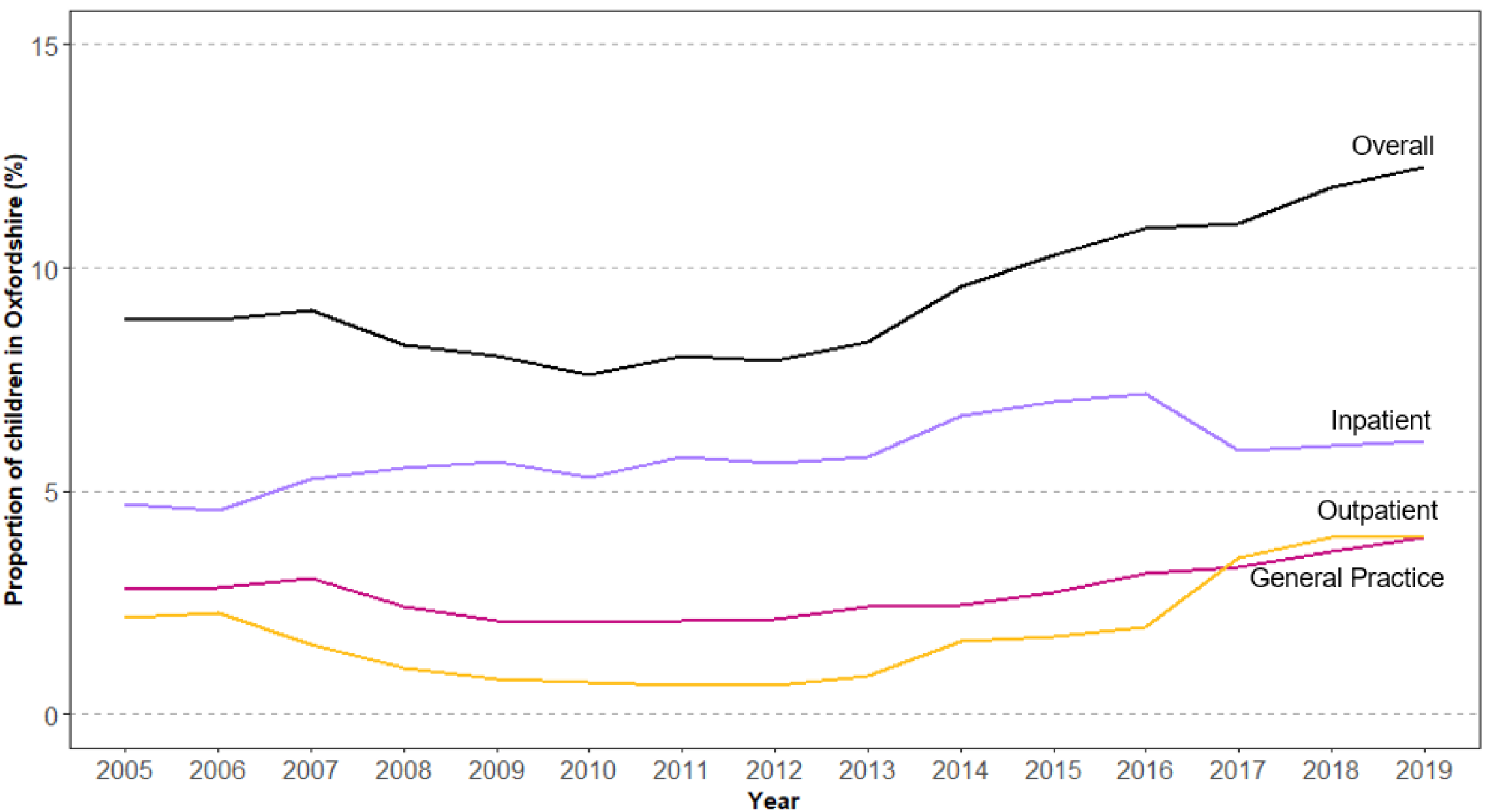
Proportion of children in Oxfordshire who had at least 1 test from 2005 to 2019 overall, and in each healthcare setting.

### Test ranking

The most frequently ordered tests from 2005 to 2019 by setting and age group are shown in **Figure 3**. The top five tests were: full blood count, urea and electrolytes, liver function tests, C-reactive protein and calcium, magnesium, phosphate levels. The top five tests remained reasonably consistent for all age groups and settings. Thyroid function tests, iron studies, IgA, coeliac screen, B12 and folate testing were most frequently requested in general practice (**Supplementary Table 2**).

**Figure 3.**
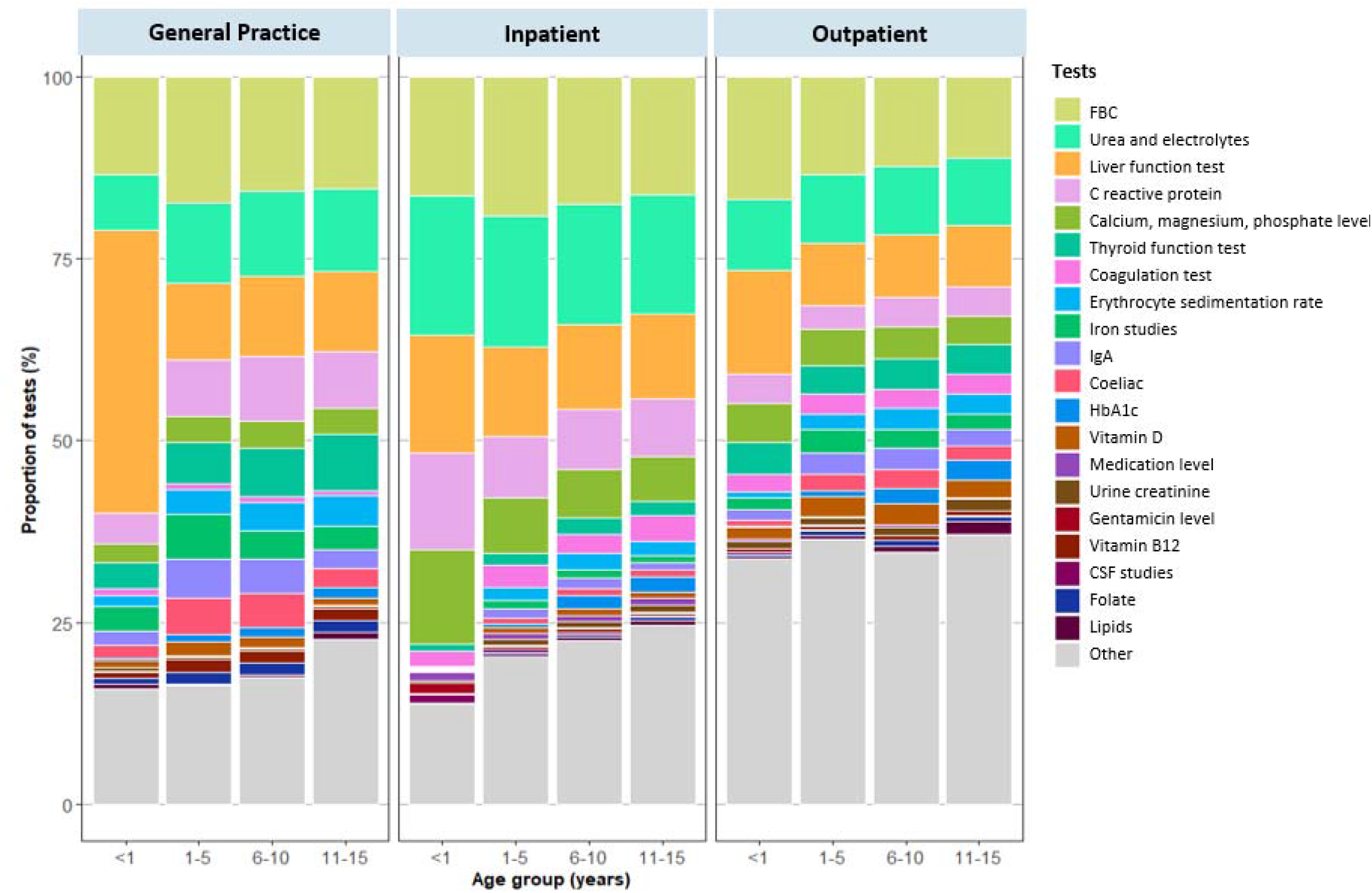
The most frequently requested tests for children in Oxfordshire from 2005 to 2019, by setting and age.

### Trends in specific test use

The temporal changes in the top 25 most frequently requested tests are shown in **Supplementary Figure 1**. Test use followed specific patterns:

- Continuous increase: coeliac testing, creatine kinase, CSF studies, folate, HbA1c, IgA, Iron studies, medication level, parathyroid hormone, thyroid function test, vitamin B12, vitamin D
- Continuous decrease: coagulation profile, gentamicin levels, monospot test for glandular fever
- Mixed: amylase, calcium-magnesium-phosphate levels, C-reactive protein, creatine kinase (CK), IgG/IgMfull blood count (FBC), glucose, lipids, liver function tests (LFT), urea and electrolytes, thyroid function tests (TFT), urine creatinine

The average annual percentage change for each test is presented in **Figure 4**. Vitamin D testing had the largest average annual change, increasing by 26·5% per year (95% CI 23·7 to 29·3%, p<0·001), followed by parathyroid hormone testing, which increased by 9·8% per year (95% CI 6·8 to 12·9%, p<0·001) and iron studies, rising by 9·3% per year (95% CI 7·3 to 11·4%, p<0·001). Other tests that demonstrated a significant increase were (in decreasing order of AAPC): Vitamin B12, folate, HbA1c, IgA levels, coeliac test, creatine kinase, CSF studies, thyroid function tests and IgG/IgM. For those tests with decreasing rates, testing for glandular fever (monospot) fell by the largest margin of 8·8% per year (95% CI -11·4 to -6·1%, p<0·001), followed by gentamicin testing, which decreased by 6·1% per year (95% CI -10·9 to -1·1%, p=0·02) and coagulation tests which declined by 2·9% per year (95% CI -6·8 to 1·2%, p=0·2).

**Figure 4.**
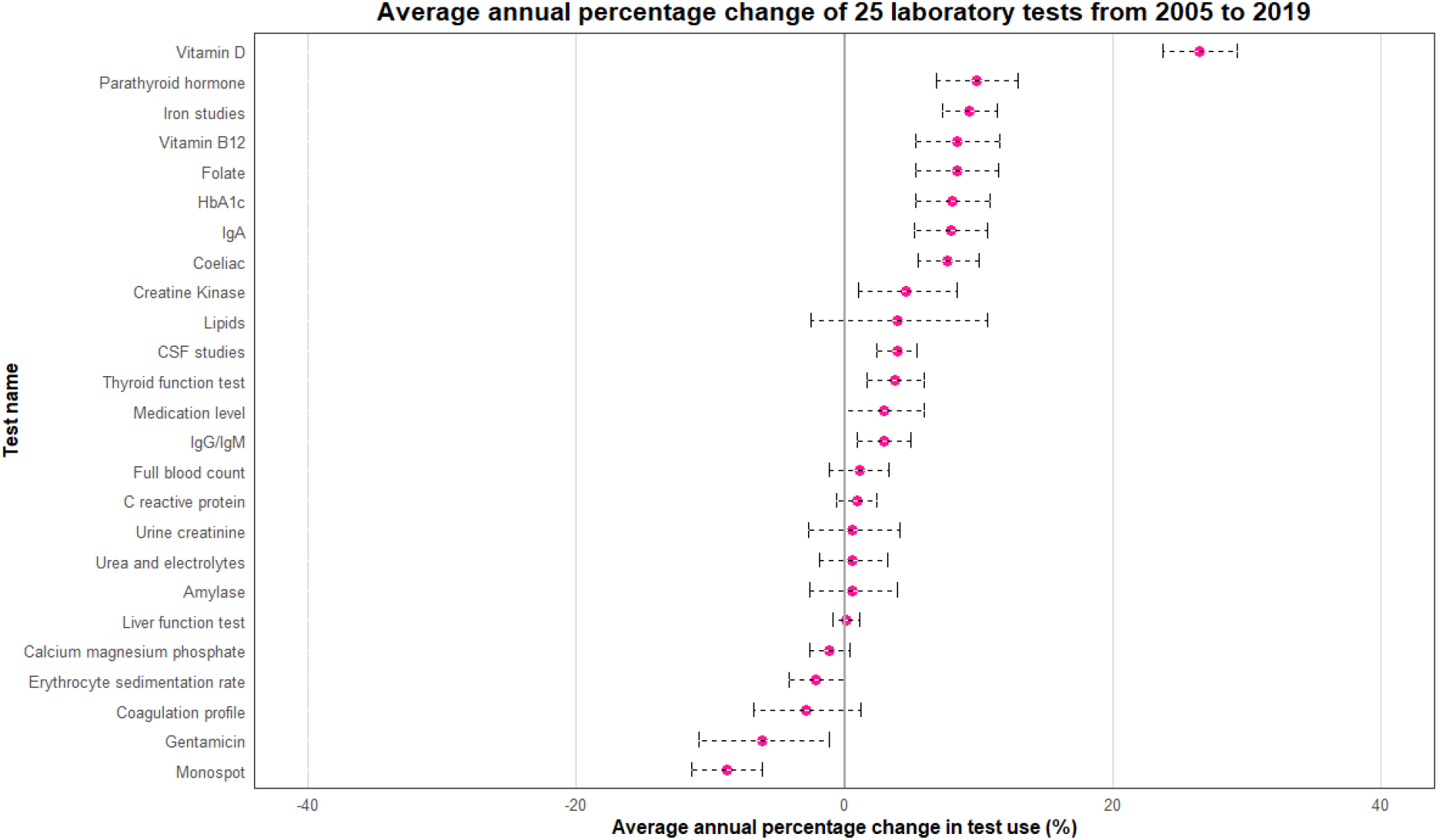
Average annual percentage change in test use for 25 specific tests from 2005 to 2019.

### Changes in test use by age and setting

When stratified by age group (**Supplementary Figure 2**) and setting (**Supplementary Figure 3**), testing increased consistently for vitamin D. For other tests, trends were not uniformly distributed across ages and settings. For example, parathyroid hormone levels, iron studies, folate and vitamin B12 testing increased in the 1–15-year-olds, more so in general practice compared with other settings. HbA1c testing increased in general practice and inpatient settings but decreased in the outpatient setting. CRP testing significantly increased in general practice from 2011, with an annual percentage increase of 9% per year after this point (95% CI 5·1% to 13·0%, p<0·001).

## Discussion

In this descriptive study, we identified trends in test use for children from 2005 to 2019 in Oxfordshire by sex, age and setting. Our results demonstrated that after an initial decline, testing rates for children increased from 2012 to 2019. This change was more pronounced in females than males. The largest relative increases occurred in the outpatient setting. The only age group that experienced a decline in testing rates over the study period was infants under the age of one, who predominantly had tests performed in hospital. Fewer tests were likely conducted for admitted children, as NHS Digital data estimates an increase in admission rates for children aged less than 1 year to the Oxford University Hospitals (OUH) NHS Trust over the same period. ^13,14^

Of the most common tests, testing for vitamin D, parathyroid hormone, iron studies, folate, and vitamin B12 increased by the greatest proportion annually. Relative increases in these tests were most pronounced in general practice. These are consistent with temporal changes in test use by adults in UK primary care from 2000 to 2015,^8^ where O’Sullivan and colleagues reported increases in testing for vitamin D (which increased by 54% per year), iron (16% per year), ferritin (19% per year), vitamin B12 (17% per year), folate (18% per year) and CRP (17% per year). There are several possible reasons for this. In recent years, there has been greater awareness of some conditions such as vitamin D deficiency and iron deficiency, making it more likely that doctors will test for these conditions. Increased disease prevalence may also explain the rise in testing.^15^ Iron studies, folate, B12, thyroid function tests and creatine kinase form part of the workup for fatigue, of which there is a high prevalence in children and adolescents; a 2007 prospective study of British adolescents reported the point prevalence of fatigue was 38%, but more recent estimates are lacking. Most of these tests (vitamin D, iron studies, folate, vitamin B12, coeliac test, serum IgA) are also included in investigations for malnutrition and faltering growth, suggesting increasing clinician concern and/or incidence of these conditions.

Similarly, a high prevalence of childhood constipation, with rates ranging from 5 to 30%, may explain the increased rates of testing for thyroid disorder.^16^ Increased testing for diabetes (HbA1c) and lipid profiles are likely correlated with the rising incidence of childhood obesity and metabolic syndrome, in line with 2006 NICE recommendations.^17–19^

Testing rates in Oxfordshire increased dramatically in 2012. Several possible explanations exist for this, including workforce shifts and specialty expansion. The Oxford Children’s Hospital building was opened in 2007 and offers tertiary paediatric care to children around the region; general paediatric services for Oxfordshire children are delivered within the Oxford Children Hospital and the Horton General Hospital in Banbury. The opening of the Children’s Hospital building increased both inpatient and outpatient numbers for children requiring tertiary level care. The hospital offers most paediatric medical and surgical specialties with few exceptions (including paediatric nephrology and paediatric cardiac surgery). Children who require other subspecialty services are referred to other centres such as Great Ormond Street Children’s Hospital, or Southampton Children’s Hospital. Over time, the tertiary specialty workload has increased, with an increased number of referrals from all around the region outside of Oxfordshire (including Berkshire, Buckinghamshire, Wiltshire, Milton Keynes). The most pronounced increases in testing occurred in the outpatient setting which expanded during the study period. According to NHS digital data, from 2009 to 2020, the number of children seen in OUH outpatient clinics increased by 63% (with a sharp increase in 2013).^20^ We could not determine which tests were conducted among Oxfordshire residents compared to referrals for tests from other areas, or which were for specialty or general paediatric patients. As a result, the appropriateness of the denominator may have changed over time, influencing the observed trends. General paediatric services for Oxfordshire across the two sites underwent a major service change in 2011, with eleven new general paediatric Consultants appointed. There have been several changes in the junior staffing structure, numbers, and experience over the years (also experienced by other Trusts around the country); junior staff care for both general and specialty patients. Whilst all staff are appropriately qualified and experience for their post, in general some members of staff, at all levels, may have less experience than their peers fifteen years go. Less experienced doctors may be inclined to test more,^21,22^ which may be one explanation for the increase in testing rates. In addition, parental expectations and anxiety levels have increased, and more experienced doctors are more likely to try to manage expectations by explanation, than by testing.

The rise in testing rates may represent over-testing in some instances. CRP is not recommended as routine blood testing in general practice, and its rise in this setting likely represents inappropriate use. ^23^ In the case of vitamin D testing, NICE guidance only recommends testing in children if they have musculoskeletal symptoms, abnormal serum bone profile or X-Ray findings, suspected bone disease such as osteomalacia or known bone disease such as osteoporosis.^24^ A retrospective analysis of vitamin D testing across all age-groups in the Northumbria Healthcare NHS Trust from 2002 to 2017 found a similar rise in testing rates.

Over three-quarters of the tests performed in those aged below 30 were found to have an inappropriate clinical indication.^25^ If many vitamin D tests return with insufficiency or deficiency, it may be worthwhile to recommend routine vitamin D supplementation for children rather than routine blood testing.

Increased parental anxiety and expectation as potential drivers of over-testing may also be relevant to primary care. There has been a decline in the numbers of experienced family doctors who are more likely to reassure parents without testing and referring children, and a concurrent rise in less experienced GPs.^26^ Children who are referred by these GPs likely present to outpatient appointments with parents expecting their child to undergo a diagnostic workup.

Our study is the first to describe long-term trends in test use in a population-based study of children. Test variation over time indicates potential changes in disease prevalence but also suggests potentially inappropriate testing. In addition, we identified areas of potential overuse, including CRP testing in general practice, and vitamin D testing.

The generalisability of our findings beyond Oxfordshire is unclear. Oxfordshire, on average, is a less socially deprived region with high educational attainment, and our results of increased test use over time may reflect greater access to laboratory tests in this area. However, other regions of the UK may show different trends and/or lower testing rates. If this is the case, it may support a hypothesis of over-testing in Oxfordshire.

We excluded glucose and blood gas tests because we thought they could be unreliably coded in the laboratory data (in some cases these are performed as bedside point-of-care care tests rather than being sent to the laboratory for analysis); however, these were the 8th and 15th most common tests, based on the potentially under-reported data. Estimations of temporal trends in test use and the proportion of children receiving at least one test, therefore, may be generalizable to laboratory tests but not point-of-care testing.

We did not have individual patient-level data or patient records containing complete information on the clinical indications for each test. This would have been beneficial as it would have provided insight into the appropriateness of tests. For example, it would be advantageous to compare whether children who have complex conditions and frequently visit the hospital (e.g., for cystic fibrosis pulmonary optimization) are more likely to have blood tests than others. If patient-level data were available, sensitivity analyses excluding children with high testing rates, would potentially provide a more accurate view of trends in testing for the general population.

The findings of this descriptive analysis suggest some areas of potentially inappropriate test use, but this needs to be confirmed by comparing the testing rates with the corresponding test results (i.e., testing rates increasing with decreasing rates of abnormal results). In saying this, the thresholds for diagnosing a result as ‘abnormal’ are not consistently adjusted for sex and age, leading to children being misdiagnosed with conditions that may represent normal. The pediatric CALIPER study set out to establish reference ranges for a wide range of biochemical markers using different analytic methods.^27^ Therefore, further analyses using this dataset should consider paediatric reference intervals with age and sex-adjusted thresholds.

We limited testing data until the end of 2019 to eliminate the impacts of the COVID-19 pandemic. However, further research should be conducted to explore the pandemic’s impacts. A systematic review examining changes in global healthcare use during the pandemic, demonstrated a 31% decline in diagnostic imaging and tests, based on 12 included studies.^28^ The pandemic serves as a natural experiment, allowing us to examine the impacts of decreased healthcare utilization. Reduced testing almost certainly resulted in missed diagnoses and deleterious outcomes in some cases, but the authors of this review found that reductions often tended to be greater for milder or less severe illness, suggesting that in some cases, forgoing tests resulted in reduced harm from unnecessary testing. The review did not find any primary studies conducted in children, and future research focusing on how the use of childhood diagnostics changed during the pandemic may determine which tests, if any, could be considered unnecessary.

Laboratory tests do not account for all the tests that children receive. Further analyses of testing variation should include other tests, including urine testing, microbiology and infection, imaging, and spirometry. The findings of this study can be compared with other settings to examine if the changes in test use are consistent across England and in other places with similar paediatric healthcare systems. Larger datasets including individual patient-level data and demographics could be used to determine if testing rates are linked to deprivation levels and ethnicity.

The analyses we have presented allow clinicians to become aware of their test-ordering practices. Increased test use exacerbates the burden on physicians with increased patient workload and time pressures. It also puts considerable strain on health expenditure. While testing is crucial in certain situations, every clinician should consider if a test is likely to yield more benefit than harm to the child, their family and the overall health system.

## Conclusions

Laboratory test use by children in Oxfordshire has increased overall since 2005, especially within the outpatient setting. Vitamin D tests have increased by the highest margin, which reflects greater clinician awareness but potential overuse. Further research should examine trends using other diagnostics and compare trends with other regions to determine if geographic variation exists.

## Data Availability

All R code used for data management, analysis and creating the figures is archived online at https://github.com/elizabethtthomas/paediatric-testing-oxfordshire.

## Acknowledgements

The authors would like to thank Dr Janet Craze for her clinical insights and comments on this manuscript, and Dr Patrick Fahr for assistance with statistical analysis.

## Author contributions

ETT contributed to study conceptualisation, methodology, completed the data analyses and wrote the original draft. DRW reviewed the statistical aspects of the study and provided critical input on the original manuscript. BS extracted and verified the data, contributed to data interpretation and reviewed the draft manuscript. PG provided supervisory input and comments and feedback on the manuscript. RP provided supervisory input, contributed to conceptualisation, methodology, reviewed the statistical aspects of the study and revision of the manuscript. CH also provided supervisory input, and contributed to conceptualisation, methodology, data interpretation and revision of the manuscript. All authors had full access to all the data in the study and had final responsibility for the decision to submit for publication.

## Data sharing statement

The study protocol, statistical analysis plan and analytic code is openly available. The study protocol was preregistered on the Open Science Framework (OSF: https://doi.org/10.17605/OSF.IO/KE6DM). All R code used for data management, analysis and creating the figures is archived online at https://github.com/elizabethtthomas/paediatric-testing-oxfordshire.

## Supplementary Figures and Tables

**Supplementary Table 1.** Number of tests requested by age group and setting

|  |  |  |  |
| --- | --- | --- | --- |
| Age group | Setting | Number of children | % |
| <1 year | Total | 580,636 | 100.0 |
|  | General Practice | 4,157 | 0.7 |
|  | Inpatient | 558,716 | 96.2 |
|  | Outpatient | 17,763 | 3.1 |
| 1-5 years | Total | 439,770 | 100.0 |
|  | General Practice | 49,123 | 11.2 |
|  | Inpatient | 315,321 | 71.7 |
|  | Outpatient | 75,326 | 17.1 |
| 6-10 years | Total | 319,387 | 100.0 |
|  | General Practice | 85,081 | 26.6 |
|  | Inpatient | 170,158 | 53.3 |
|  | Outpatient | 64,148 | 20.1 |
| 11-15 years | Total | 409,632 | 100.0 |
|  | General Practice | 155,145 | 37.9 |
|  | Inpatient | 188,361 | 46.0 |
|  | Outpatient | 66,126 | 16.1 |

**Supplementary Table 2.** Top 25 requested tests from 2005 to 2019, stratified by setting

| Test | Total | General Practice |  | Inpatient |  | Outpatient |  |
| --- | --- | --- | --- | --- | --- | --- | --- |
|  | n | n | % | n | % | n | % |
| Full blood count | 321,421 | 53,288 | 16.6 | 234,446 | 72.9 | 33,687 | 10.5 |
| Urea and electrolytes | 305,959 | 38,420 | 12.6 | 242,947 | 79.4 | 24,592 | 8.0 |
| Liver function tests | 250,612 | 38,183 | 15.2 | 188,658 | 75.3 | 23,771 | 9.5 |
| C reactive protein | 180,448 | 27,342 | 15.2 | 143,092 | 79.3 | 10,014 | 5.5 |
| Calcium magnesium phosphate | 154,806 | 11,842 | 7.6 | 131,150 | 84.7 | 11,814 | 7.6 |
| Thyroid function tests | 54,322 | 23,950 | 44.1 | 19,627 | 36.1 | 10,745 | 19.8 |
| Coagulation tests | 45,291 | 2,451 | 5.4 | 35,769 | 79.0 | 7,071 | 15.6 |
| Erythrocyte sedimentation rate | 35,200 | 13,271 | 37.7 | 15,515 | 44.1 | 6,414 | 18.2 |
| Iron studies | 29,576 | 13,283 | 44.9 | 9,541 | 32.3 | 6,752 | 22.8 |
| IgA | 28,838 | 12,335 | 42.8 | 9,656 | 33.5 | 6,847 | 23.7 |
| Coeliac | 24,804 | 11,890 | 47.9 | 7,309 | 29.5 | 5,605 | 22.6 |
| HbA1c | 18,769 | 4,620 | 24.6 | 9,543 | 50.8 | 4,606 | 24.5 |
| Vitamin D | 18,435 | 4,331 | 23.5 | 7,266 | 39.4 | 6,838 | 37.1 |
| Medication level | 14,401 | 301 | 2.1 | 13,516 | 93.9 | 584 | 4.1 |
| Urine creatinine | 12,998 | 1,040 | 8.0 | 8,742 | 67.3 | 3,216 | 24.7 |
| Gentamicin level | 10,629 | 19 | 0.2 | 10,560 | 99.4 | 50 | 0.5 |
| Vitamin B12 | 10,478 | 5,570 | 53.2 | 3,193 | 30.5 | 1,715 | 16.4 |
| CSF studies | 9,835 | 68 | 0.7 | 9,469 | 96.3 | 298 | 3.0 |
| Folate | 9,825 | 5,281 | 53.8 | 2,849 | 29.0 | 1,695 | 17.3 |
| Lipids | 9,676 | 2,388 | 24.7 | 4,682 | 48.4 | 2,606 | 26.9 |
| Amylase | 8,108 | 640 | 7.9 | 6,614 | 81.6 | 854 | 10.5 |
| IgG/IgM | 7,992 | 1,135 | 14.2 | 4,455 | 55.7 | 2,402 | 30.1 |
| Creatine Kinase | 7,236 | 557 | 7.7 | 4,480 | 61.9 | 2,199 | 30.4 |
| Parathyroid hormone | 6,739 | 282 | 4.2 | 4,122 | 61.2 | 2,335 | 34.6 |
| Monospot test | 6,214 | 4,670 | 75.2 | 1,346 | 21.7 | 198 | 3.2 |

**Supplementary Figure 1.**
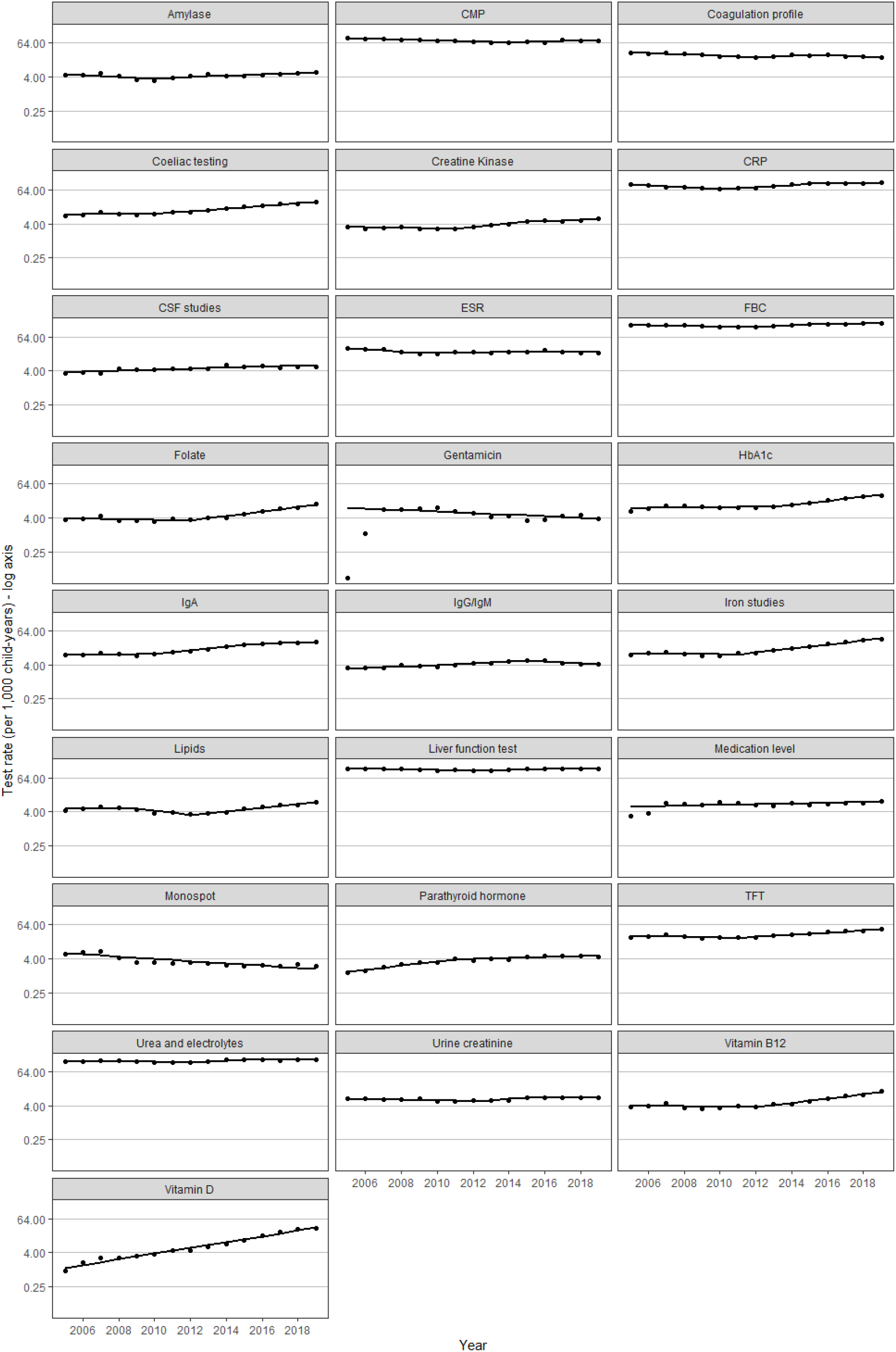
Temporal patterns in test use for the top 25 tests by children in Oxfordshire, 2005 to 2019.

**Supplementary Figure 2.**
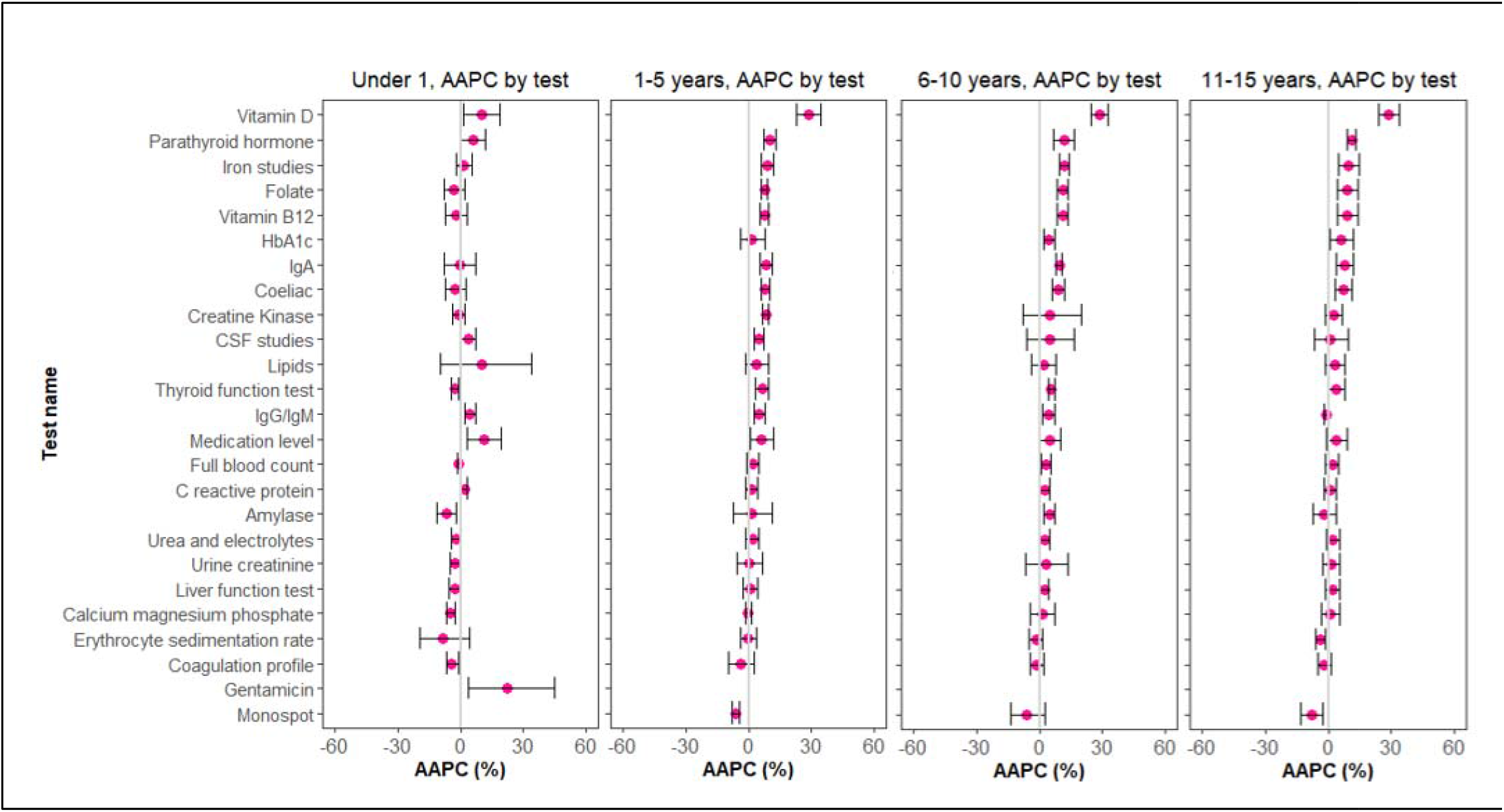
Average annual percentage change in specific test use by age in Oxfordshire, 2005 to 2019. **Footnote** We excluded tests from the joinpoint analysis if there were fewer than 5 tests performed in a given year, hence there are missing AAPC data points for some tests (HbA1c, monospot, gentamicin).

**Supplementary Figure 3.**
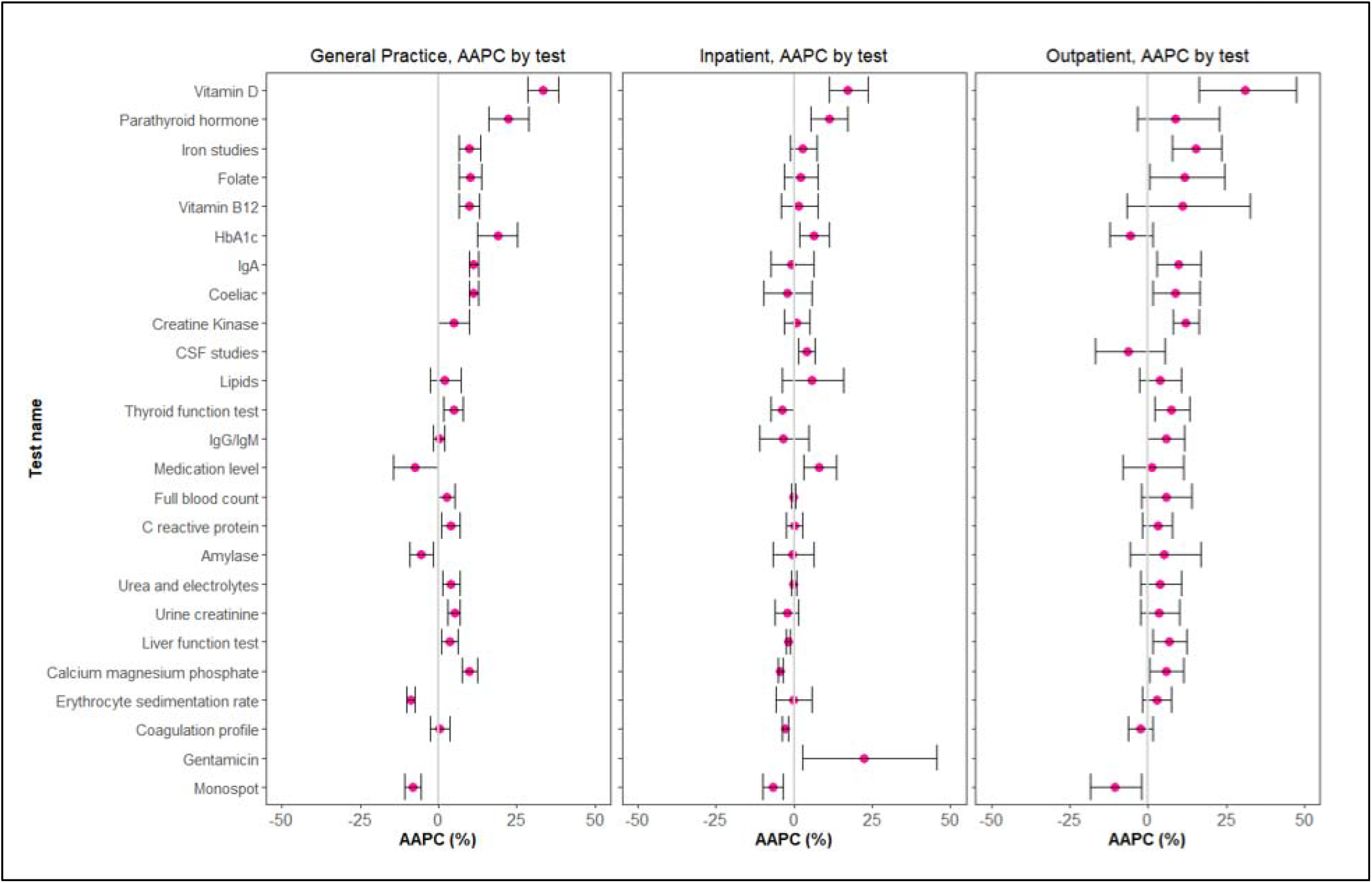
Average annual percentage change in specific test use by setting in Oxfordshire, 2005 to 2019. **Footnote** We excluded tests from the joinpoint analysis if there were fewer than 5 tests performed in a given year, hence there are missing AAPC data points for some tests (CSF studies, gentamicin).

